# Descriptive epidemiology of 16,780 hospitalized COVID-19 patients in the United States

**DOI:** 10.1101/2020.07.17.20156265

**Authors:** Devika Chawla, Shemra Rizzo, Kelly Zalocusky, Daniel Keebler, Jenny Chia, Lisa Lindsay, Vincent Yau, Tripthi Kamath, Larry Tsai

**Affiliations:** Genentech, Inc., 1 DNA way, South San Francisco, California 94080, USA

## Abstract

1.

**Background:** Despite the significant morbidity and mortality caused by the 2019 novel coronavirus disease (COVID-19), our understanding of basic disease epidemiology remains limited. This study aimed to describe key patient characteristics, comorbidities, treatments, and outcomes of a large U.S.-based cohort of patients hospitalized with COVD-19 using electronic health records (EHR).

**Methods:** We identified patients in the Optum De-identified COVID-19 EHR database who had laboratory-confirmed COVID-19 or a presumptive diagnosis between 20 February 2020 and 6 June 2020. We included hospitalizations that occurred 7 days prior to, or within 21 days after, COVID-19 diagnosis. Among hospitalized patients we describe the following: vital statistics and laboratory results on admission, relevant comorbidities (using diagnostic, procedural, and revenue codes), medications (NDC, HCPC codes), ventilation, intensive care unit (ICU) stay, length of stay (LOS), and mortality.

**Findings:** We identified 76,819 patients diagnosed with COVID-19, 16,780 of whom met inclusion criteria for COVID-related hospitalization. Over half the cohort was over age 50 (74.5%), overweight or obese (77.2%), or had hypertension (56.6%). At admission, 30.3% of patients presented with fever (>38° C) and 32.3% had low oxygen saturation (<90%). Among the 16,099 patients with complete hospital records, we observed that 58.9% had hypoxia, 23.4% had an ICU stay during hospitalization, 18.1% were ventilated, and 16.2% died. The median LOS was 6 days (IQR: 4, 11).

**Interpretation:** To our knowledge, this is the largest descriptive study of patients hospitalized with COVID-19 in the United States. We report summary statistics of key clinical outcomes that provide insights to better understand COVID-19 disease epidemiology.

## 2. INTRODUCTION

The novel coronavirus disease (COVID-19) poses an unprecedented public health challenge with significant morbidity and mortality. In March 2020, the World Health Organization (WHO) officially declared COVID-19 a pandemic. As of 30 July 2020, the number of deaths attributed to the virus numbered over 140,000 in the United States, and these numbers continue to rise every day.^1^

Patients with COVID-19 present with a wide range of symptoms, including but not limited to fever, cough, shortness of breath, headache or body aches, fatigue, and/or loss of taste and smell.^2,3^ Transmission primarily occurs during person-to-person contact via respiratory droplets, but can potentially occur by touching a surface with the virus and subsequently touching your eyes, mouth, or nose via fomites.^4^ Additionally, many reports suggest asymptomatic cases and transmission, which pose a unique challenge to epidemiological studies of this disease.^5^

The scientific community is quickly gaining insights into the SARS-COV-2 virus and clinical spectrum of COVID-19 disease, and rigorous epidemiological studies from real-world settings are a key component to fully understanding this novel disease. Specifically, detailed epidemiologic studies of hospitalized patients—which reflect a more severe subset of the infected population—can help inform scientific understanding of disease progression, comorbidities, treatment patterns, and important clinical endpoints like ventilation and ICU stay. While some real-world studies of hospitalized patients in the United States have been published,^6,7,8,9^ few studies possess the necessary sample size and inpatient medication details to provide a robust snapshot of patients with COVID-19. Additionally, reproducing results from various study populations is necessary for synthesis and understanding of the true nature of the disease.^10^

To help fill this knowledge gap, the primary objective of this study is to describe key patient characteristics, comorbidities, treatments, and outcomes in a large U.S.-based cohort of patients hospitalized with COVID-19 using electronic health records (EHR). Additionally, the study describes vital signs and clinical conditions upon admission. To the authors’ knowledge, this study describes the largest cohort of patients hospitalized with COVID-19 in the United States to date.

## 3. METHODS

To address this research objective, individuals with COVID-19 were extracted from the Optum® De-identified COVID-19 EHR dataset from 20 February 2020 to 6 June 2020, sourced from a dataset containing patient-level medical and administrative records from hospitals, emergency departments, outpatient centers, and laboratories from across the United States. Data are de-identified in compliance with the HIPAA Expert Method and managed according to Optum® customer data use agreements. The COVID-19 EHR dataset sources clinical information from hospital networks that provide data meeting Optum’s internal data quality criteria.

Two cohorts were created; an overall COVID-19 cohort, and a subset of this cohort, a hospitalized cohort. To be eligible for the overall COVID-19 cohort, patients required at least one of the following: a diagnosis code of U07.1, U07.2, a positive diagnostic test for COVID-19, or a diagnosis code of B97.29 with the absence of a negative molecular SARS-CoV-2 test within a 14-day window. Index date was set to be the first instance of a laboratory-confirmed or presumptive COVID-19 diagnosis. Sensitivity analyses were run on the cohort definition (see Supplementary Material).

To be eligible for the hospitalized COVID-19 cohort, patients additionally had to meet at least one of the following criteria: (1) inpatient or emergency room (ER) overnight visit with initial COVID-19 diagnosis occurring during hospitalization and within 7 days of hospital admission, or (2) inpatient or ER overnight visit within 21 days of initial COVID-19 diagnosis date, where hospitalization has record of COVID-19 diagnosis. A hospitalization was defined as inpatient or ER visits that included at least one overnight stay. Contiguous ER and inpatient visits, with up to a 1-day gap, were considered a single hospitalization. In this study, we capture information from the first COVID-19 hospitalization.

Missing end dates from inpatient records were imputed using evidence of continuous inpatient presence using diagnostic, administered medications, and vital records, allowing for a 1-day gap. The last date of continuous presence was used as the hospitalization end date. This imputation method was validated using inpatient records with known end dates as well as by physician review.

As the Optum EHR data included patient records for individuals who may still be hospitalized at the time when the database was censored, patient records were treated the following way: (1) patients who remained in-hospital at the time of data delivery were included in estimations of COVID-related outcomes and of clinical characteristics on admission, and (2) patients with incomplete hospitalization up to a week before data delivery were excluded from estimates that require completed hospitalization (length of stay [LOS], intensive care unit [ICU], ventilation, medications administered).

Patient characteristics for the overall and hospitalized COVID-19 cohorts are reported in Tables 1 and 2. Comorbidities were assessed using relevant ICD-9/10 diagnosis codes in the 1 year prior to index date. We report the presence of any pregnancy-related diagnosis code in the 6 months prior to index date. The Charlson Comorbidity Index (CCI) reported in Table 2 is a summary metric for describing comorbidity burden in patients.^13^

**Table 1.** Patient demographics among all COVID-19 cases (N=76,819) and a subset of hospitalized COVID-19 cases (N=16,780)

| Demographics |  | Missing | Overall | Hospitalized |
| --- | --- | --- | --- | --- |
| **n** |  |  | 76,819 | 16,780 |
| **Age<sup>a</sup>, median [Q1,Q3]** |  | 2 (0·0) | 54·0 [38·0,67·0] | 62·0 [49·0,75·0] |
| **Age category, n (%)** | <18 |  | 1,767 (2·3) | 190 (1·1) |
|  | 18-29 |  | 8,214 (10·7) | 802 (4·8) |
|  | 30-39 |  | 11,034 (14·4) | 1,351 (8·1) |
|  | 40-49 |  | 11,635 (15·1) | 1,933 (11·5) |
|  | 50-64 |  | 21,606 (28·1) | 4,906 (29·2) |
|  | 65-74 |  | 10,946 (14·2) | 3,298 (19·7) |
|  | 75-84 |  | 6,689 (8·7) | 2,537 (15·1) |
|  | >85 |  | 4,926 (6·4) | 1,763 (10·5) |
| **Gender, n (%)** | Female | 45 (0·06) | 42,362 (55·2) | 7,953 (47·4) |
|  | Male |  | 34,412 (44·8) | 8,822 (52·6) |
| **Race, n (%)** | African American | 17,636 (23·0) | 16,236 (27·4) | 4,667 (35·1) |
|  | Asian |  | 2,399 (4·1) | 464 (3·5) |
|  | Caucasian |  | 40,548 (68·5) | 8,150 (61·4) |
| **Ethnicity, n (%)** | Hispanic | 14,681 (19·1) | 9,368 (15·1) | 2,184 (15·0) |
|  | Not Hispanic |  | 52,770 (84·9) | 12,402 (85·0) |
| **Region, n (%)** | Midwest | 5,744 (7·5) | 30,212 (42·5) | 7,312 (46·0) |
|  | Northeast |  | 36,093 (50·8) | 7,806 (49·1) |
|  | South |  | 666 (0·9) | 57 (0·4) |
|  | West |  | 4,104 (5·8) | 733 (4·6) |
| **Division, n (%)** | East North Central | 2,010 (2·6) | 22,201 (29·7) | 5,633 (34·5) |
|  | East South Central |  | 666 (0·9) | 57 (0·3) |
|  | Middle Atlantic |  | 28,880 (38·6) | 5,894 (36·0) |
|  | Mountain |  | 1,006 (1·3) | 178 (1·1) |
|  | New England |  | 7,213 (9·6) | 1,912 (11·7) |
|  | Pacific |  | 3,098 (4·1) | 555 (3·4) |
|  | **South Atlantic/West South Central** |  | 3,734 (5·0) | 443 (2·7) |
|  | **West North Central** |  | 8,011 (10·7) | 1,679 (10·3) |
| **Smoking, n (%)** | **Current smoker** | 16,935 (22·0) | 5,185 (8·7) | 1,254 (8·6) |
|  | **Never smoked** |  | 38,735 (64·7) | 8,693 (59·8) |
|  | **Previously smoked** |  | 15,964 (26·7) | 4,585 (31·6) |
| **Insurance, n (%)** | **Commercial** | 7,119 (9·3) | 37,508 (53·8) | 6,084 (37·3) |
|  | **Medicaid** |  | 8,619 (12·4) | 2,410 (14·8) |
|  | **Medicare** |  | 16,553 (23·7) | 5,972 (36·6) |
|  | **Other** |  | 6,257 (9·0) | 1,659 (10·2) |
|  | **Uninsured** |  | 763 (1·1) | 178 (1·1) |
| **BMI category, n (%)** | **Underweight (<18.5)** | 19,839 (25·8) | 1,446 (2·5) | 404 (2·6) |
|  | **Normal (18.5 to <25)** |  | 12,520 (22·0) | 3,168 (20·2) |
|  | **Overweight (25 to <30)** |  | 17,135 (30·1) | 4,601 (29·4) |
|  | **Obese (>30)** |  | 25,879 (45·4) | 7,493 (47·8) |
| **Recently pregnant<sup>b</sup>, n (%)** | **No** | 0 (0·0) | 75,475 (98·3) | 16,447 (98·0) |
|  | **Yes** |  | 1,344 (1·7) | 333 (2·0) |
| **Hospitalizations, n (%)** | **Hospitalization started before COVID-19 diagnosis** | 0 (0·0) | 1,594 (2·1) | 1,594 (9·5) |
|  | **Hospitalization within 14 days** |  | 15,104 (19·7) | 15,104 (90·0) |
|  | **Hospitalized within 14 to 21 days** |  | 82 (0·1) | 82 (0·5) |
|  | **Not hospitalized** |  | 60,039 (78·2) |  |

<sup>a</sup> The Optum EHR database aggregates ages of those 89 and older.
<sup>b</sup> Recently pregnant was defined as the presence of pregnancy-related ICD 9/10 diagnosis codes in the 6 months prior to COVID-19 diagnosis date.

**Table 2.**
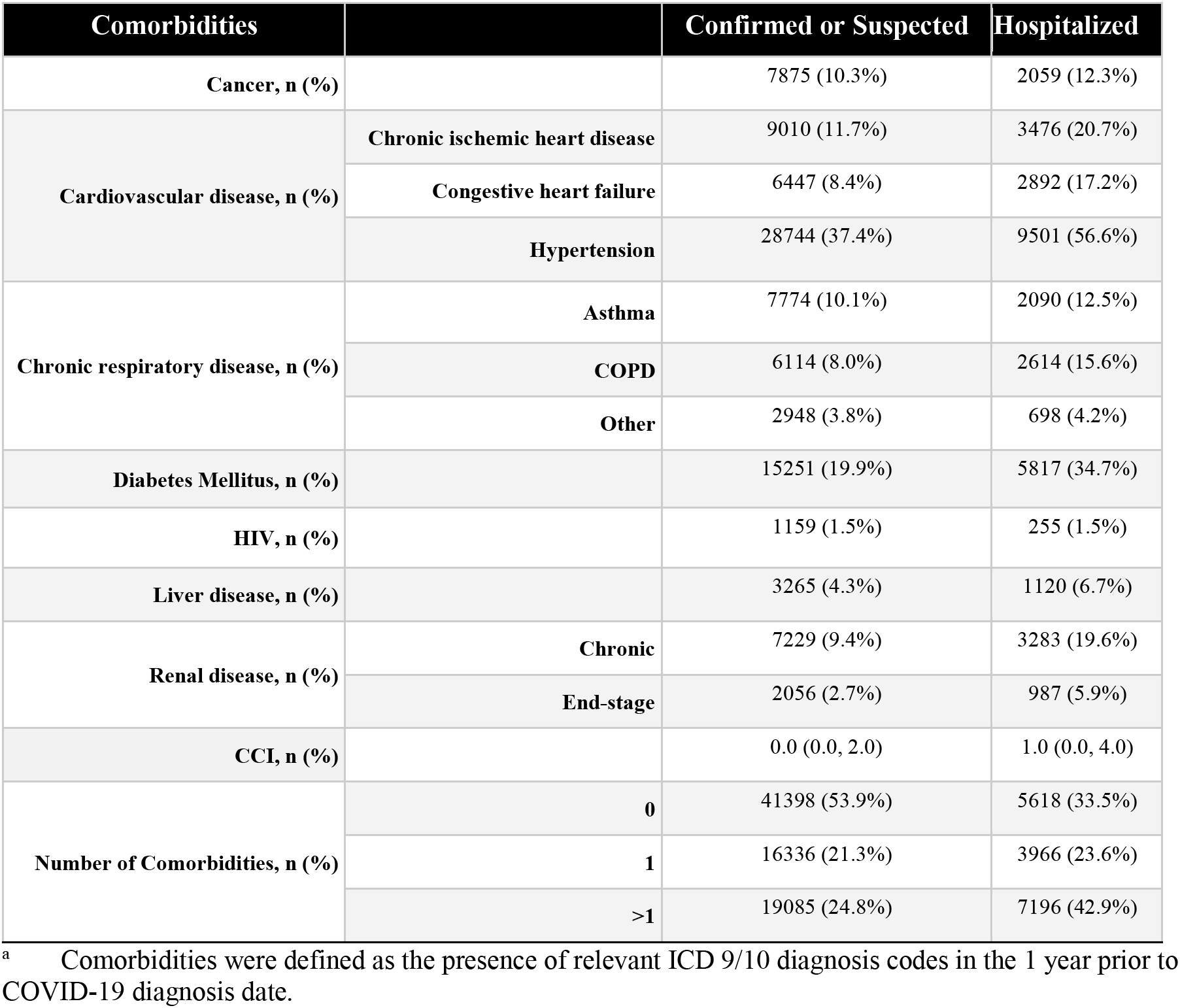
Patient comorbidities among all COVID-19 cases (N=76,819) and a subset of hospitalized COVID-19 cases (N=16,780)

Selection of relevant clinical outcomes, laboratory values, and medications was based on prior publications and/or clinical consultation. Clinical conditions and outcomes for the patients hospitalized with COVID-19 are reported in Table 3, overall and age-stratified. Hypoxia, any mechanical ventilation, invasive ventilation, bilevel positive airway pressure (BiPAP) or continuous positive airway pressure (CPAP) therapy, and extracorporeal membrane oxygenation (ECMO) were identified using ICD-9/10 diagnosis, ICD-9/10 PCS, CPT, and REV codes. Respiratory failure including acute respiratory distress syndrome (ARDS), pneumonia, and sepsis were identified using ICD-9/10 diagnosis codes, and reported stratified by timing of occurrence.

**Table 3.** Key clinical conditions and outcomes among hospitalized COVID-19 cases (N=16,780), stratified by age, N (%)

| **Outcomes on Admission<sup>a</sup>** |  | **All hospitalized** | **<18** | **18-65** | **>65** |
| --- | --- | --- | --- | --- | --- |
| **n** |  | 16,780 | 190 | 9,334 | 7,256 |
| **Respiratory failure on admission** |  | 6923 (41·3) | 33 (17·4) | 3,407 (36·5) | 3,483 (48·0) |
| **Pneumonia on admission** |  | 10,190 (60·7) | 19 (10·0) | 5,571 (59·7) | 4,600 (63·4) |
|  | **Bacterial pneumonia** | 611 (3·6) | 2 (1·1) | 314 (3·4) | 295 (4·1) |
| **Sepsis on admission** |  | 3,416 (20·4) | 16 (8·4) | 1,695 (18·2) | 1,705 (23·5) |
| **Outcomes after Admission and Before Discharge<sup>b</sup>** |  | **All hospitalized** | **<18** | **18-65** | **>65** |
| **n<sup>a</sup>** |  | 16,099 | 185 | 8,958 | 6,956 |
| **Respiratory failure after admission** |  | 1887 (11·7) | 8 (4·3) | 929 (10·4) | 950 (13·7) |
| **Pneumonia after admission** |  | 1,622 (10·1) | 5 (2·7) | 879 (9·8) | 738 (10·6) |
|  | **Bacterial pneumonia** | 313 (1·9) | 2 (1·1) | 156 (1·7) | 155 (2·2) |
| **Sepsis after admission** |  | 942 (5·9) | 8 (4·3) | 458 (5·1) | 476 (6·8) |
| **Outcomes Anytime During Hospitalization** |  | **All hospitalized** | **<18** | **18-65** | **>65** |
| **n<sup>a</sup>** |  | 16,099 | 185 | 8,958 | 6,956 |
| **Mortality<sup>c</sup>** |  | 2,613 (16·2) | 2 (1·1) | 560 (6·3) | 2,051 (29·5) |
| **Hospitalization LOS, median [Q1,Q3]** |  | 6·0 [4·0,11·0] | 3·0 [2·0,5·0] | 6·0 [3·0,9·0] | 7·0 [5·0,12·0] |
| **Hypoxia** |  | 9,478 (58·9) | 35 (18·9) | 4,794 (53·5) | 4,649 (66·8) |
| **Ventilation** |  | 2,919 (18·1) | 14 (7·6) | 1,348 (15·0) | 1,557 (22·4) |
|  | **CPAP/BiPAP<sup>c</sup>** | 671 (4·2) | 7 (3·8) | 289 (3·2) | 375(5·4) |
|  | **Invasive ventilation** | 2,085 (13·0) | 6 (3·2) | 973 (10·9) | 1,106 (15·9) |
| **ICU** |  | 3,768 (23·4) | 31 (16·8) | 1,777 (19·8) | 1,960 (28·2) |
|  | **ICU LOS, median [Q1, Q4]** | 4·0 [1·0,10·0] | 2·0 [1·0,7·0] | 4·0 [1·0,10·0] | 4·0 [1·0,10·0] |

|  | Hospitalization LOS<sup>d</sup>,<br>median [Q1, Q4] | 11.0 [7.0,19.0] | 5.0 [3.0,9.5] | 12.0 [7.0,19.0] | 11.0 [7.0,18.0] |
| --- | --- | --- | --- | --- | --- |
| ECMO<sup>e</sup> |  | 38 (0.2) | 0 (0.0) | 35 (0.4) | 3 (0.0) |

<sup>a</sup> Conditions measured throughout or at the end of hospitalization were reported only among completed hospitalizations (N=16,099), while conditions measured on admission were reported among all hospitalizations (N=16,780).
<sup>b</sup> Outcomes after admission but before discharge capture day 2 onwards of hospitalization.
<sup>c</sup> Mortality reflects inpatient death within study period.
<sup>d</sup> Hospitalization LOS reflects the total LOS, including ICU stay, for patients who had any ICU stay.
<sup>e</sup> ECMO: Extracorporeal membrane oxygenation; CPAP: continuous positive airway pressure; BiPAP: bilevel positive airway pressure

Mortality was captured primarily from EHR records where death has been documented. Only month and year of death were available; exact day of death was not included in the dataset.

Select laboratory values for the hospitalized cohort are reported in Table 5. We report the first measurement of each analyte within a 3-day window of hospital admission. We exclude laboratory values with non-numeric results or missing unit values. For troponin, we additionally exclude laboratory values with missing reference ranges since this laboratory value is reported in the present study as a proportion above normal range. If multiple testing methods were reported for the same analyte, the most prevalent test(s) were used until greater than 95% of measurements were captured. Similarly, when multiple units were reported for the same analyte, the most prevalent unit(s) were used, and units were harmonized where appropriate.

Relevant medications administered during hospitalization are reported in Table 6.^14,15,16,17^ When appropriate, medication classes from ATC were converted to NDC or HCPCS, and these NDC and HCPCS codes were used to ascertain medications administered. Since remdesivir lacked an NDC code at the time of this analysis, we used a free-text match for ‘remdesivir’ in the medication descriptions. Due to particular clinical interest in these drugs, we report dexamethasone separately from other corticosteroids; similarly, we report azithromycin separately from other antibiotics.

As the objectives of this analysis were descriptive; no statistical significance testing was conducted. Python was used to conduct all analyses. ^18,19,20^

## 4. RESULTS

We identified 76,819 patients that met the inclusion criteria for the overall COVID-19 cohort. Of these, 51.8% had a positive diagnostic test, 76.2% had a U07.1 diagnosis, and 16.2% had a B97.29 diagnosis. We observed very few pediatric patients (younger than 18 years of age) (2.3%), while 29.4% of the cohort was above the age of 65. Among those with race/ethnicity data, the cohort was 27.4% African-American and 15.1% Hispanic, though race/ethnicity data were missing for almost a quarter of patients. Most patients came from the Northeast (50.8%) or Midwest (42.5%). More than half of patients were overweight or obese (75.5%, among patients with non-missing values), though BMI was missing in 25.8% of patients (see Table 1).

Within the broader cohort, we then identified 16,780 patients (21.8%) that met inclusion criteria for the hospitalized COVID-19 cohort. Of these, 16,099 had a completed hospitalization. We again observed very few pediatric patients (1.1%), while 45.2% of this cohort was over the age of 65. Over three-quarters of the hospitalized patients were overweight or obese (77.2%, among patients with non-missing values), which was consistent with the overall COVID-19 cohort. Similar patterns for race, ethnicity, and geographic region were observed as compared to the overall cohort (see Table 1). African-American patients had a higher rate of hospitalization as compared to white patients (28.7% vs. 20.0%). Approximately 16.2% of patients with completed COVID-19 hospitalizations had death recorded between hospital admission and end of study period (see Table 3).

Comorbidity burden was high in the overall and hospitalized cohorts. In particular, diabetes mellitus (19.9%, 34.7%) and hypertension (37.4%, 56.6%) were prevalent in the overall and hospitalized cohorts, respectively. Approximately 53.9% of the overall cohort had no reported CCI-eligible comorbidities, as compared to 33.5% of the hospitalized cohort (see Table 2). A sensitivity analysis to confirm patient presence in the EHR system indicated that patients lacking records prior to COVID-19 index date accounted for <0.01% (approximately 1 in 20,000 patients), indicating low missing data in this 1-year comorbidity lookback. Those with at least one CCI-eligible comorbidity were hospitalized at a higher rate than those without comorbidities (31.5% vs. 13.6%).

We observed that 58.9% of patients had hypoxia at any point during hospitalization. Among the hospitalized cohort, 18.1% of patients were ventilated, 4.2% had non-invasive CPAP or BiPAP therapy, 13.0% had invasive ventilation, and 23.4% of patients were in the ICU at some point during their hospitalization. Median LOS was 6 days (IQR: 4, 11), and increased with age (see Table 3). Among patients who had an ICU stay at some point during their hospitalization, the overall LOS was 11 days (IQR: 7, 19), and the ICU-specific LOS was 4 days (IQR: 1, 10). Many patients were admitted to the hospital with complications, including respiratory failure (41.3%), pneumonia (60.7%), and sepsis (20.4%).

Vital signs and laboratory values at hospital admission are reported in Tables 4 and 5. Approximately 19.8% of patients had a moderate fever (temperature of >38° C and <=39° C), and 10.5% had a high fever (temperature >39° C). The majority of patients (44.8%) had oxygen saturation between 90% and 95%, and 32.3% of patients had <90% oxygen saturation. Of note, we observed a median ferritin of 5110 ng/ml (IQR: 225.0, 1088.0), median c-reactive protein (CRP) of 793 mg/l (IQR: 34 0, 142 0), and a median fibrin d-dimer of 760.0 (IQR: 430, 1600), although missingness was considerable for the following laboratory values: creatine kinase, fibrin d-dimer, troponin, and procalcitonin.

**Table 4.** Select vitals results on admission among hospitalized COVID-19 cases (N=16,780)

| Vitals |  | Missing | N (%) |
| --- | --- | --- | --- |
| **n** |  |  | 16,780 |
| **Temperature** |  | 550 (3.3) |  |
|  | **Median [Q1, Q3]** |  | 37.4 [36.9,38.2] |
|  | **38-39° C** |  | 3,212 (19.8) |
|  | **> 39° C** |  | 1,704 (10.5) |
| **Oxygen saturation** |  | 888 (5.3) |  |
|  | **Median [Q1, Q3]** |  | 93.0 [89.0, 95.0] |
|  | **<70%** |  | 404 (2.5) |
|  | **70-80%** |  | 622 (3.9) |
|  | **80-90%** |  | 4,111 (25.9) |
|  | **90-95%** |  | 7,125 (44.8) |
|  | **>95%** |  | 3,630 (22.8) |
| **Heart rate (HR)** |  | 772 (4.6) |  |
|  | **Median [Q1, Q3]** |  | 102.0 [90.0,115.0] |
|  | **HR > 100 beats/min on admission** |  | 8,496 (52.3) |
| **Respiratory rate** |  | 668 (4.0) |  |
|  | **Median [Q1, Q3]** |  | 23.0 [20.0,29.0] |
|  | **RR >24 breaths/min on admission** |  | 6,554 (40.7) |

**Table 5.**
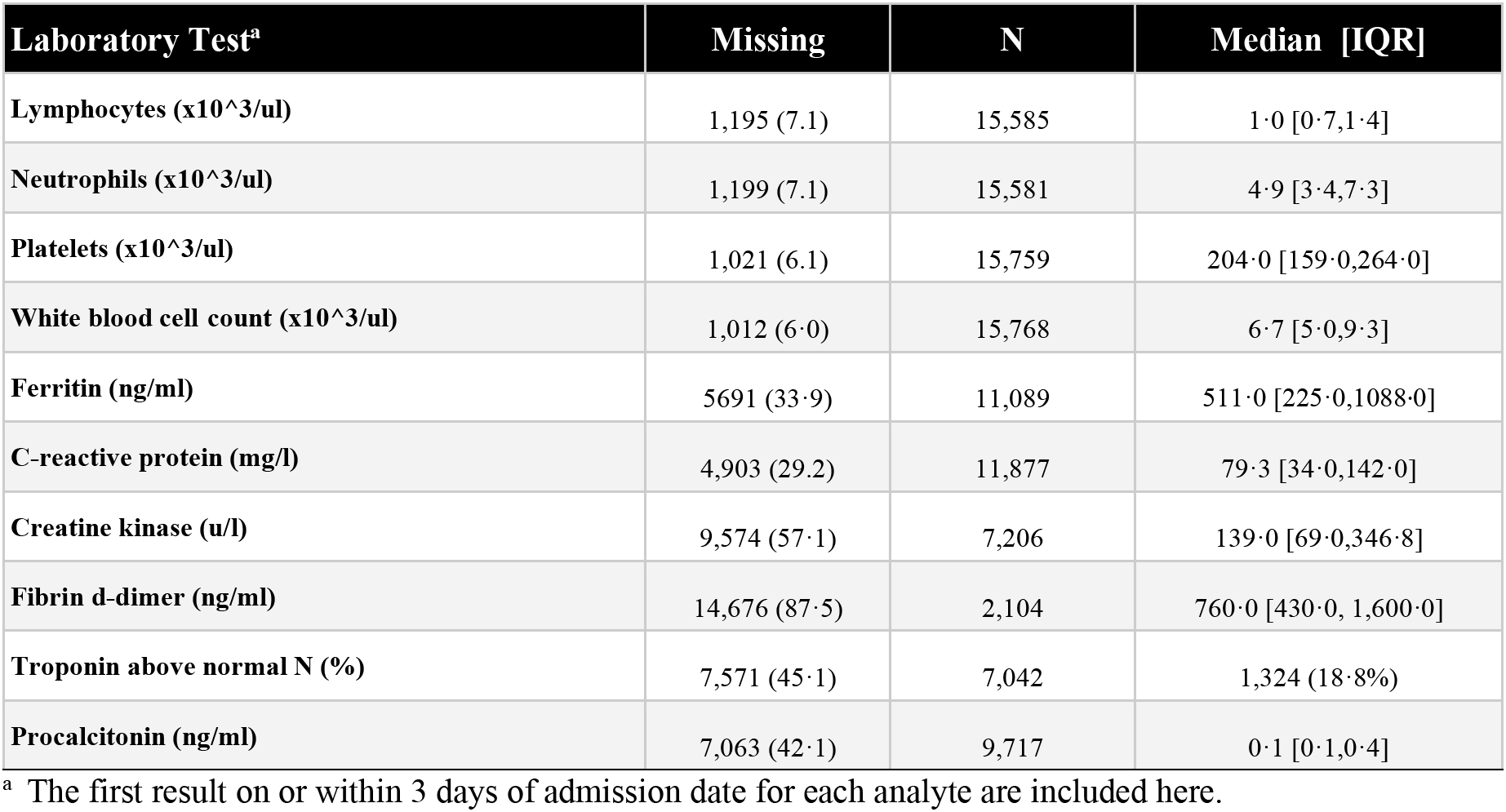
Select laboratory results on admission among hospitalized COVID-19 cases (N=16,780)

Relevant medications administered during inpatient stay are summarized in Table 6. Among the 16,099 patients with completed hospitalizations, we observed low use of tocilizumab (2.9%) and remdesivir (13%), and prevalent use of corticosteroids excluding dexamethasone (249%), dexamethasone (2.9%), hydroxychloroquine/chloroquine (35.2%), azithromycin (38.1%), antibiotics other than azithromycin (60.9%), and anticoagulants (70 9%).

**Table 6.**
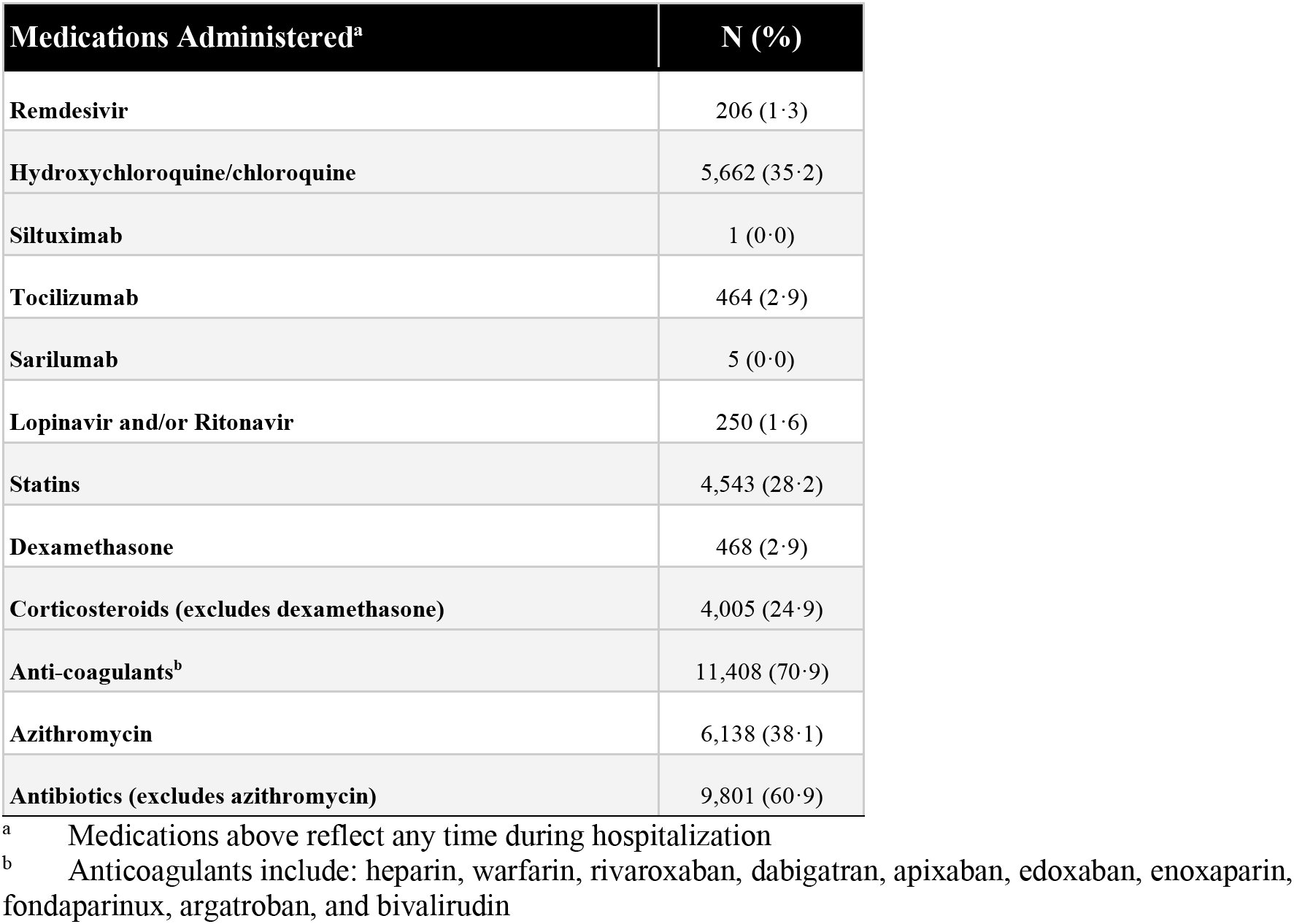
Medications administered during hospitalization among completed COVID-19 hospitalizations (N=16,099)

## 5. DISCUSSION

In this study, we report key patient characteristics and clinical outcomes in a large, multicenter cohort of patients hospitalized with COVID-19. To our knowledge, this is the largest cohort of hospitalized COVID-19 patients in the published literature. We took advantage of the richness of this EHR database to report on a wide range of clinically-relevant factors, including vitals and laboratory values on admission, medications administered, hospitalization rates, ventilation rates, and mortality. Of the 76,819 patients with COVID-19 identified in the database, we observed a hospitalization rate of 21.8%. Among the 16,099 patients with completed hospitalizations for COVID-19 in this database, we observed a mortality rate of 16.2%.

To contextualize the mortality and ICU rates observed in our hospitalized cohort, we compared our results to two recent publications: a hospitalized cohort from New York City (N=5,700, but mortality is reported on N=2,634) by Richardson et al, and a publication on patients from the Kaiser Permanente Northern California population (N=1,299, but mortality is reported on N=321) by Myers et al.^6,7^ We observed a mortality rate among hospitalized patients (16.2%) that was consistent with Myers et al (15.6%), but slightly lower than Richardson et al (21.0%). Moreover, we observed an ICU rate (23.4%) lower than Myers et al (30.0%) but higher than Richardson et al (14.2%). Finally, we observed an invasive ventilation rate (13.0%) similar to Richardson et al (12.2%) but lower than Myers et al (29.2%). Overall, our key endpoints were consistent with contemporary publications, but our slightly lower ventilation rate may reflect differences in underlying disease severity or geographic differences in clinical practice.

One strength of this study was the inclusion of vital sign records and laboratory test results, which provide an extra layer of granularity in describing the clinical epidemiology of these patients. A total of 30.3% of patients presented with fever (>38° C) at admission, which is consistent with previous reports (30.7%; Richardson et al). In contrast, we report a higher percent (32.3%) of patients presenting with low (<90%) oxygen saturation, elevated heart rate (52.3%), and elevated respiratory rate (40.7%) at admission than do Richardson et al (20.4%, 43.1%, and 17.3%). These differences could represent more severe respiratory compromise at the time of admission or could reflect differences in supplemental oxygen administration at the time of assessment. Laboratory results generally suggest less severe inflammation on admission, with median ferritin mildly elevated at 511.0 ng/ml, CRP moderately elevated at 79.3 mg/L, and fibrin d-dimer moderately elevated at 760.0 ng/ml compared with 798 ng.ml, 130 mg/l, and 438 ng/ml in Richardson et al. Median lymphocyte count was at the low end of normal but was relatively higher and neutrophil count lower at 10 and 49 x 10^3^/ul, vs. 0.88 and 5 3 x 10^3^/ul in Richardson et al, also consistent with relatively milder disease on admission. Similar proportions of patients had elevated troponin (18.8% vs. 22.6%), suggesting similar rates of cardiac involvement. Median procalcitonin was normal in both studies, consistent with viral infection.

Our analysis differed from other publications in three ways: (1) Richardson et al and Myers et al restricted their analyses to patients with laboratory-confirmed COVID-19, whereas we included patients who were either laboratory-confirmed or presumptively diagnosed, (2) we report medications administered among hospitalized patients, (3) our study describes a much larger sample of patients hospitalized with COVID-19. Richardson et al captured patients from the NYC region, Myers et al captures patients from Northern California, and our analysis captures patients across the United States but oversamples from the Northeast and Midwest regions.

Consistent with other publications, we observed significant comorbidity burden among hospitalized patients, as evidenced by the high proportions for both individual and combined (>1) comorbidities as measured by the CCI. In fact, those with at least one CCI-eligible comorbidity were hospitalized at a higher rate than those without comorbidities. We also observed a higher median age among hospitalized patients compared to the overall COVID-19 cohort. Rates of hospitalization increased by age, with 11.1% of adults younger than 40 years and 37.0% of patients older than 75 years being hospitalized. These observations are consistent with reports that older patients and patients with high comorbidity burden are more likely to develop severe disease that requires hospitalization.^21,22^

The high proportion of hospitalized patients with comorbid hypertension (56.6%) is particularly striking. This report is consistent with Richardson et al, which also reported that 56.6% of their hospitalized cohort had hypertension.^6^ Some published reports hypothesize that high rates of hypertension in patients with COVID-19 may be explained by the fact that hypertensive patients are often treated with angiotensin-converting enzyme (ACE) inhibitors, which may lead to increased expression of ACE2, increasing the risk of COVID-19 infection. ^23,24,25,26^

Pneumonia is often part of the COVID-19 sequelae.^27,28^ Using diagnosis codes, we observed pneumonia in 60·7% of patients on admission and an additional 10.1% after admission. Acute respiratory distress syndrome (ARDS) is an acute inflammatory lung condition that sometimes occurs as part of the sequelae for patients with severe COVID-19. However, ARDS is often underreported by clinicians,^29^ and instead potentially captured more broadly as respiratory failure. Hence, we report all respiratory failure, which accounted for 41.3% of patients on admission and an additional 11.7% after admission (Table 3). Bacterial pneumonia is also hypothesized to occur as a complication of COVID-19, though reports are inconsistent.^30,31,32^ While we observed bacterial pneumonia (3.6%) on admission, these results should be interpreted with caution as it may be underreported in patients who are already diagnosed with COVID-19. Furthermore, much of the bacterial pneumonia in this population may be associated with invasive ventilation, and thus develops over the course of inpatient stay rather than on admission.

We assessed medication use among patients hospitalized with COVID-19 that have been hypothesized in the literature ^14,15,16,17^ to be related to COVID-19 disease trajectories. Among the medications captured, anticoagulants were strikingly prevalent, with 70.9% of patients receiving anticoagulants at some point during hospitalization. We hypothesize that in most cases these medications were most likely used prophylactically to prevent deep vein thrombosis (DVT) due to reports of higher risk of blood clots in COVID-19 patients.^33,34^ We observed low use of remdesivir and tocilizumab (1.3% and 2.9%, respectively). Finally, we observed approximately a third of hospitalized patients received hydroxychloroquine/chloroquine, which is striking considering more recent reports on the lack of efficacy of this drug class for COVID-19.^35,36^

## 6. LIMITATIONS

This study contributes meaningfully to our understanding of the descriptive epidemiology of COVID-19 in a real-world U.S.-based setting, including disease presentation, clinical outcomes, and medication use; however, some important limitations must be considered.

First, the case definition utilized in this study relied on either a diagnosis code or positive diagnostic test. The diagnostic test aspect of the case definition allows for high specificity but potentially low sensitivity. Conversely, the diagnostic code aspect of the case definition allows for a broader capture of patients hospitalized with COVID-19 that do not have a recorded diagnostic test; this may result in lower specificity and higher sensitivity. This definition excludes a subset of patients who (a) did not get tested, (b) got tested outside of the relevant window, (c) did not receive the appropriate diagnosis code, or (d) had COVID-19, with or without signs and symptoms, but did not seek care. This may result in potential selection bias whereby our cohort reflects a more severe COVID-19 population; therefore, our hospitalization and mortality rates in the overall COVID-19 cohort should be interpreted with caution. Notably, our analysis had a broader case definition as compared to other published reports of patients hospitalized with COVID-19, which restrict to laboratory-confirmed cases.^6,7^ We conducted a sensitivity analysis removing the B97.29 code from our case definition and observed an overall cohort size reduction of 4.8% and no meaningful changes to key outcomes. Finally, it is possible that patients received a SARS-CoV-2 test within the Optum EHR system, but are admitted elsewhere, causing an underestimation of hospitalization rate in this cohort.

Given the use of EHR data that has been captured in real-time during a pandemic setting, the issue of missing data is an important consideration and impacts a number of key study outcomes. In particular, we were unable to report supplemental oxygen and ventilator-associated pneumonia due to insufficient capture of these outcomes. Additionally, we rely on diagnostic, procedural, and revenue codes to capture key conditions like pneumonia, sepsis, and respiratory failure, and outcomes such as ventilation, meaning that these features are likely to be underreported. It is also noteworthy to highlight that our mortality estimates may underestimate the true mortality rate, since we do not capture deaths that occur outside of hospitalization.

Finally, our study population may not perfectly generalize to the broader population of patients hospitalized with COVID-19. Of note, most of our cohort is from the Northeast or the Midwest. Public health response, availability of testing, social-distancing policies, hospital and ICU capacity, and discharge practices differ by hospital and geography, which leads to (a) substantial heterogeneity within our cohort, and (b) limited ability to generalize findings to the overall population of the United States.

## Data Availability

Data for this study was licensed from Optum.

## 9. SUPPLEMENTARY MATERIAL

### A. Cohort definition

The U07.1 and U07.2 ICD-10 diagnosis codes reflect the WHO and CDC guidelines for coding confirmed or probable COVID-19 cases effective date 1 April 2020 ^37,38,39^.To ensure full capture of the earliest COVID-19 cases, we also included the B97.29 code (‘Other coronavirus as the cause of disease classified elsewhere’), which was the interim code used before the U07.1 and U07.2 codes were implemented on 1 April 2020 ^40,41^.

For the diagnostic laboratory tests used for inclusion, we only allowed COVID-19 tests that explicitly have the following strings in their name: ‘pcr’, ‘rna’, ‘naat’, ‘np’ (nasopharyngeal), or ‘op’ (oral-pharyngeal) to ensure tests were diagnostic tests; antibody tests were not used as part of our case definition. For the index date used in Tables 1 and 2, earliest date of diagnosis code or positive diagnostic test was used. We did not impose any age restrictions on the study population.

A sensitivity analysis was performed to evaluate the inclusion of the 4.8% of cases that had a B97.29 code and no U07.1 diagnosis or positive lab. No changes were observed in the results.

Additionally, a sensitivity analysis was performed to evaluate the inclusion of COVID-19 diagnosis codes in the definition. No meaningful changes were observed in the results when restricting to a cohort of patients with a positive PCR laboratory test (n=39,798).

## References

1. Centers for Disease Control and Prevention – Daily Updates of Totals by Week and State. [Updated 10 July 2020; Accessed on 17 July 2020]. Available from https://www.cdc.gov/nchs/nvss/vsrr/COVID19/index.htm

2. Centers for Disease Control and Prevention – Symptoms of Corona Virus. [Updated 13 May 2020; Accessed on 17 July 2020]. Available from: https://www.cdc.gov/coronavirus/2019-ncov/symptoms-testing/symptoms.html

3. Guan WJ, Zheng-Yi N, Hu Y, et al. Clinical Characteristics of Coronavirus Disease 2019 in China. N Engl J Med 2020;382:1708–20.

4. Centers for Disease Control and Prevention – How Covid-19 Spreads. [Updated 16 June 2020; Accessed 17 July 2020]. Available from: https://www.cdc.gov/coronavirus/2019-ncov/prevent-getting-sick/how-covid-spreads.html

5. Gandhi M, Yokoe DS, Havlir DV. Asymptomatic Transmission, the Achilles’ Heel of Current Strategies to Control Covid-19. N Engl J Med 2020;382:2158–60.

6. Richardson S, Hirsch JS, Narasimhan M, et al. Presenting Characteristics, Comorbidities, and Outcomes Among 5700 Patients Hospitalized With COVID-19 in the New York City Area. JAMA 2020;323:2052–59.

7. Myers LC, Parodi SM, Escobar GJ, Liu VX. Characteristics of Hospitalized Adults with COVID-19 in an Integrated Health Care System in California. JAMA 2020;323:2195–98.

8. Cummings MJ, Baldwin MR, Abrams D, et al. Epidemiology, clinical course, and outcomes of critically ill adults with COVID-19 in New York City: a prospective cohort study. Lancet 2020; 395:1763–70.

9. Paranjpe I, Russak A, De Freitas JK, et al. Clinical Characteristics of Hospitalized Covd-19 Patients in New York City. medRxiv [Posted 26 April 2020; Accessed on 17 July 2020]. Available from: https://doi.org/10.1101/2020.04.19.20062117

10. Lipsitch M, Swerdlow DL, Finelli L. Defining the Epidemiology of Covid-19 – Studies Needed. N Engl J Med 2020;385:1194–96.

11. Halpern R, Seare J, Tong J, et al. Using Electronic Health Records to Estimate the Prevalence of Agitation in Alzheimer disease/dementia. Int J Geriatr Psychiatry 2019;34:420–31.

12. Nunes AP, Yang J, Radican L, et al. Assessing Occurrence of Hypoglycemia and Its Severity From Electronic Health Records of Patients With Type 2 Diabetes Mellitus. Diabetes Res Clin Pract 2016;121:192–203.

13. Glasheen WP, Cordier T, Gumpina R, Haugh G, Davis J, Renda A. Charlson Comorbidity Index: ICD-9 Update and ICD-10 Translation. Am Health Drug Benefits 2019;12:188–97.

14. Pascarella G, Strumia A, Piliego C, et al. COVID-19 Diagnosis and Management: A Comprehensive Review. J Intern Med 2020; epub ahead of print doi: 10.1111/joim.13091.

15. Guo YR, Cao QD, Hong ZS, et al. The Origin, Transmission and Clinical Therapies on Coronavirus Disease 2019 (COVID-19) Outbreak – An Update on the Status. Mil Med Res 2020;7:11.

16. Johnson RM, Vinetz JM. Dexamethasone in the Management of Covid -19. BMJ 2020;370: doi: 10.1136/bmj.m2648.

17. Rossi B, Nguyen LS, Zimmermann P, et al. Effect of tocilizumab in hospitalized patients with severe pneumonia COVID-19: a cohort study. medRxIV 2020; posted 9 June 2020. Available from: https://doi.org/10.1101/2020.06.06.20122341

18. The pandas development team. (2020, March 18). pandas-dev/pandas: Pandas 1.0.1 (Version v1.0.1). Zenodo. http://doi.org/10.5281/zenodo.3715232

19. McKinney W. Data structures for statistical computing in python. In Proceedings of the 9th Python in Science Conference 2010;445:51–6.

20. Oliphant TE. A guide to NumPy. Vol. 1. Trelgol Publishing USA;2006.

21. Centers for Disease Control and Prevention – Preliminary Estimates of the Prevalence of Selected Underlying Health Conditions Among Patients with Coronavirus Disease 2019 — United States, February 12–March 28, 2020. Weekly 2020;69:382–6.

22. Zheng Z, Peng F, Xu B, et al. Risk Factors of Critical & Mortal COVID-19 Cases: A Systematic Literature Review and Meta-Analysis. J Infect 2020;S0163-4453(20)30234-6. doi: 10.1016/j.jinf.2020.04.021. online ahead of print.

23. Fang L, Karakiulakis G, Roth M. Are Patients With Hypertension and Diabetes Mellitus at Increased Risk for COVID-19 Infection? Lancet Respir Med 2020;8:e21. doi: 10.1016/S2213-2600(20)30116-8.

24. Lo KB, McCullough PA, Rangaswami J. Antihypertensive Drugs and Risk of COVID-19? Lancet Respir Med 2020;8:e29. doi: 10.1016/S2213-2600(20)30156-9.

25. Yan R, Zhang Y, Li Y, Xia L, Guo Y, Zhou Q. Structural Basis for the Recognition of SARS-CoV-2 by Full-Length Human ACE2. Science 2020;367:1444–8.

26. Dauchet L, Lambert M, Gauthier V, et al. ACE inhibitors, AT1 receptor blockers and COVID-19: clinical epidemiology evidences for a continuation of treatments. The ACER-COVID study. medRxIV posted 1 May 2020.

27. Ahn D, Shin H, Kim M. Current Status of Epidemiology, Diagnosis, Therapeutics, and Vaccines for Novel Coronavirus Disease 2019 (COVID-19). J Microbiol Biotechnol. 2020;30:313–24.

28. Center for Disease Control and Prevention. Interim Clinical Guidance for Management of Patients with Confirmed Coronavirus Disease (COVID-19). [Updated 30 June 2020; Accessed 17 July 2020]. Available from: https://www.cdc.gov/coronavirus/2019-ncov/hcp/clinical-guidance-management-patients.html

29. Rezoagli E, Fumagalli R, Bellani G. Definition and Epidemiology of Acute Respiratory Distress Syndrome. Ann Transl Med 2017;5:282.

30. Rawson TM, Moore LSP, Zhu N, et al. Bacterial and Fungal Co-Infection in Individuals With Coronavirus: A Rapid Review to Support COVID-19 Antimicrobial Prescribing. Clin Infect Dis 2020 May 2;ciaa530. doi: 10.1093/cid/ciaa530. Online ahead of print.

31. Hughes S, Troise O, Donaldson H, Mughal N, Moore LSP. Bacterial and Fungal Coinfection Among Hospitalised Patients With COVID-19: A Retrospective Cohort Study in a UK Secondary Care Setting. Clin Microbiol Infect 2020;S1198-743X(20)30369-4. doi: 10.1016/j.cmi.2020.06.025. Online ahead of print.

32. Wu CP, Adhi F, Highland K. Recognition and Management of Respiratory Coinfection and Secondary Bacterial Pneumonia in Patients with COVID-19. Cleve Clin J Med 2020;ccc015. doi: 10.3949/ccjm.87a.ccc015. Online ahead of print.

33. Demelo-Rodriguez P, Cervilla-Munoz E, Ordieres-Ortega L, et al. Incidence of Asymptomatic Deep Vein Thrombosis in Patients With COVID-19 Pneumonia and Elevated D-dimer Levels. Thromb Res 2020;192:23–26. doi: 10.1016/j.thromres.2020.05.018.

34. Connors JM, Levy JH. COVID-19 and Its Implications for Thrombosis and Anticoagulation. Blood 2020;135:2033–2040. doi: 10.1182/blood.2020006000.

35. Ferner RE, Aronson JK. Chloroquine and Hydroxychloroquine in covid-19. BMJ 2020;369:m1432. doi: 10.1136/bmj.m1432.

36. Das S, Bhowmick S, Tiwari S, Sen S. An Updated Systematic Review of the Therapeutic Role of Hydroxychloroquine in Coronavirus Disease-19 (COVID-19). Clin Drug Investig 2020;40:591–601. doi: 10.1007/s40261-020-00927-1.

37. World Health Organization. Classifications – List of Official ICD-10 updates. Available from https://www.who.int/classifications/icd/icd10updates/en/

38. World Health Organization. Classifications – COVID-19 coding in ICD-10 25 March 2020. Available from https://www.who.int/classifications/icd/COVID-19-coding-icd10.pdf

39. Centers for Disease Control and Prevention – ICD-10-CM Official Coding Guidelines – Supplement Coding encounters related to COVID-19 Coronavirus Outbreak Effective: February 20, 2020. Available from https://www.cdc.gov/nchs/data/icd/ICD-10-CM-Official-Coding-Gudance-Interim-Advice-coronavirus-feb-20-2020.pdf

40. Centers for Disease Control and Prevention – ICD-10-CM Official Coding and Reporting Guidelines April 1, 2020 through September 30, 2020. Available from https://www.cdc.gov/nchs/data/icd/COVID-19-guidelines-final.pdf

41. Nalleballe K, Onteddu SR, Sharma R, et al. Spectrum of neuropsychiatric manifestations in COVID-19. Brain, Behavior, and Immunity. 2020;88:71–74. doi: 10.1016/j.bbi.2020.06.020.

